# Operationalized releases of *w*AlbB *Wolbachia* in *Aedes aegypti* lead to sharp decreases in dengue incidence dependent on *Wolbachia* frequency

**DOI:** 10.1101/2023.11.08.23298240

**Authors:** Ary A. Hoffmann, Nazni Wasi Ahmad, Ming Keong Wan, Yoon Ling Cheong, Noor Afizah Ahmad, Nick Golding, Nicholas Tierney, Jenarun Jelip, Perada Wilson Putit, Norhayati Mokhtar, Sukhvinder Singh Sandhu, Sai Ming Lau, Khadijah Khairuddin, Kamilan Denim, Norazman Mohd Rosli, Hanipah Shahar, Topek Omar, Muhammad Kamarul Ridhuan Ghazali, Nur Zatil Aqmar Mohd Zabari, Mohd Arif Abdul Karim, Mohamad Irwan Saidin, Muhammad Nizam Mohd Nasir, Tahir Aris, Steven P Sinkins

## Abstract

In *Aedes aegypti* mosquitoes, introduction of certain strains of inherited *Wolbachia* symbionts results in transmission blocking of various viruses of public health importance, including dengue. This has resulted in a ‘replacement’ strategy for dengue control involving release of male and female mosquitoes, whereupon *Wolbachia* is able to spread through *Ae. aegypti* populations to high frequency and reduces the incidence of dengue. *Wolbachia* strain *w*AlbB is an effective transmission blocker and stable at high temperatures, making it very suitable for use in hot tropical climates. Following the first trial field releases of the *w*AlbB strain in *Ae. aegypti* in Malaysia, releases of *w*AlbB *Ae. aegypti* have for the first time become operationalized by the Malaysian health authorities. We report here on changes in dengue incidence based on a set of 20 releases sites and 76 control sites in high rise residential areas, which allows us to directly assess the impact of *Wolbachia* frequency on dengue incidence. The results indicate an average reduction in dengue of 62.4% (confidence intervals 50-71%); importantly the level of suppression increased with *Wolbachia* frequency, with suppression of 75.8% (confidence intervals 61-87%) estimated at 100% *Wolbachia* frequency. These findings emphasize the large impacts of *w*AlbB *Wolbachia* invasions on dengue incidence in an operational setting, with the expectation that the level of dengue will further decrease as wider areas are invaded.

## Introduction

Releases of *Wolbachia*-carrying *Aedes aegypti* mosquitoes were first initiated in north-eastern Australia in 2010 [1] with the aim of replacing *Wolbachia*-free populations with populations carrying a high frequency of this bacterium that can decrease the transmission of dengue and other arboviruses [e.g. 2, 3, 4]. The initial releases were undertaken with a *Wolbachia* strain (*w*Mel) sourced from *Drosophila melanogaster* [1] but since that time releases have also been initiated with other *Wolbachia* including the *w*AlbB strain originating from *Ae. albopictus*, which causes similar levels of virus transmission blockage in multiple population backgrounds [2, 3] although blockage with a different *w*AlbB variant has been found to vary between genetic backgrounds [5]. Both invasions of *w*Mel and *w*AlbB have had a substantial impact on the local incidence of dengue [6–8] although there have been issues in securing and maintaining high *Wolbachia* infection frequencies in populations in some areas [6, 9].

For *w*Mel, local transmission cycles of seasonally imported dengue have been virtually eliminated in invaded areas in Australia [7]. In Yogyakarta in Indonesia [8] where dengue is endemic there has been an overall estimated reduction of 77%. Releases of *w*AlbB in dengue hotspots that acted as initial release sites resulted in an estimated decrease in dengue of 40.3% compared to control sites around the Kuala Lumpur region [6]. More variable reductions have been obtained in sites around Rio de Janeiro where invasion has been challenging, with *Wolbachia* frequencies often failing to reach 50% in populations and averaging 32% in release areas [9, 10].

The relative success of these research interventions despite some challenges has increased interest around the world in the operationalization of *Wolbachia* releases. A range of *w*Mel and *w*AlbB *Wolbachia* strains as well as others are starting to be developed for in country use [e. g. 11, 12] and releases have been initiated in multiple locations with local resources, with cost-effectiveness analyses supporting the widescale use of this technology [13].

In Malaysia, country operations have focused on a *Wolbachia w*AlbB strain introduced into a local genetic background [14]. The Malaysian *Wolbachia* project was launched in 2017, releasing *w*AlbB-carrying *Aedes aegypti* in 11 dengue hotspots in the Klang Valley around Kuala Lumpur [6]. Since then, *Wolbachia* density and frequency as well as dengue blocking ability have remained stable in the original release areas [15, 16] and releases are now expanding into other areas.

Because of the promising effect of the initial releases in reducing dengue incidence, the Disease Control Department of the Malaysian Ministry of Health, in collaboration with the Institute of Medical Research, deployed *Wolbachia*-carrying *Ae. aegypti* to various additional dengue hotspots localities around Kuala Lumpur. These activities coupled with ongoing monitoring at some of the original release sites and long-term data on dengue incidence provide a unique opportunity to more accurately assess dengue impact associated with *w*AlbB invasion, particularly within the context of variable *Wolbachia* frequencies.

## Methods

### Operational program

The *Wolbachia* operational program started on 7 July 2019 and has been undertaken in several phases as resources and other factors such as COVID lockdown have allowed. Phase 1 of the operational programme involved releasing eggs in containers provided to the Ministry of Health field workers. Two other phases have since been added, resulting in 40 sites now where releases have been conducted. The focus here is on the first 15 operational sites representing the first two operational phases and the 5 sites from the original program [6] where sufficient *Wolbachia* and dengue data is available across 5 years to test for intervention effects.

### Mosquito culture

All aspects of rearing the mosquitoes and quality control follow the description in Nazni et al. [6] with minor modifications. Briefly, weighed eggs were submerged in seasoned tap water (after desiccation) and exposure to an air vacuum to stimulate hatching. The seasoned water consisted of tap water stored overnight to dechlorinate the water. Eggs were hatched in beakers with the seasoned water.

For mass rearing adults, mosquito eggs were laid on paper and around 15000 eggs estimated by weight were soaked in 1 l of seasoned water in a 36 x 26 x 5.5 cm container. Two days after the hatching process, the larvae were filtered and put into a beaker containing 500 ml of seasoned water and 10 aliquots of larvae (1 ml per aliquot) were taken from the beaker using a 10 ml plastic pipette (approximately 50 larvae per 1 ml aliquot). 10 of these aliquots were placed into a 36 x 26 x 5.5 cm plastic container with 1 l of seasoned water. Sera Vipan powder (Heinsberg, Germany) was given daily to ensure even pupation.

To separate sexes, 12,000 larvae and pupae were introduced into a pupal separation system (Orinno Technologies, Singapore). These immature stages were separated over a period of around 13 min, producing groups of separated (mostly) male pupae, (mostly) female pupae and larvae.

For generalizing the next generation of the stock population of mosquitoes, we used 3750 female and 1250 male pupae which were transferred to a plastic container filled with seasoned water and placed in a mosquito cage (30.5 x 30.5 x 30.5 cm) to emerge into adults. Emerged mosquitoes were fed on laboratory-reared mice before oviposition on paper to restart the process. Wing measurements were carried out periodically for quality control and wild type males collected from field stock were introduced every 7 generations to ensure fresh material entered the culture regularly.

### Operational release sites

*Wolbachia* release areas in the Selangor region (Figure 1) were based on selection criteria important to local health staff and managers which focused on the incidence of dengue, presence of mosquitoes, and perceived barriers (mostly roads) around the release sites given that most movement of mosquitoes tends to be within buildings [c. f. 17]. The majority of the research sites, and thirteen operational release sites were situated in Selangor, while five sites were in Kuala Lumpur and two sites were in Putrajaya.

**Figure 1.**
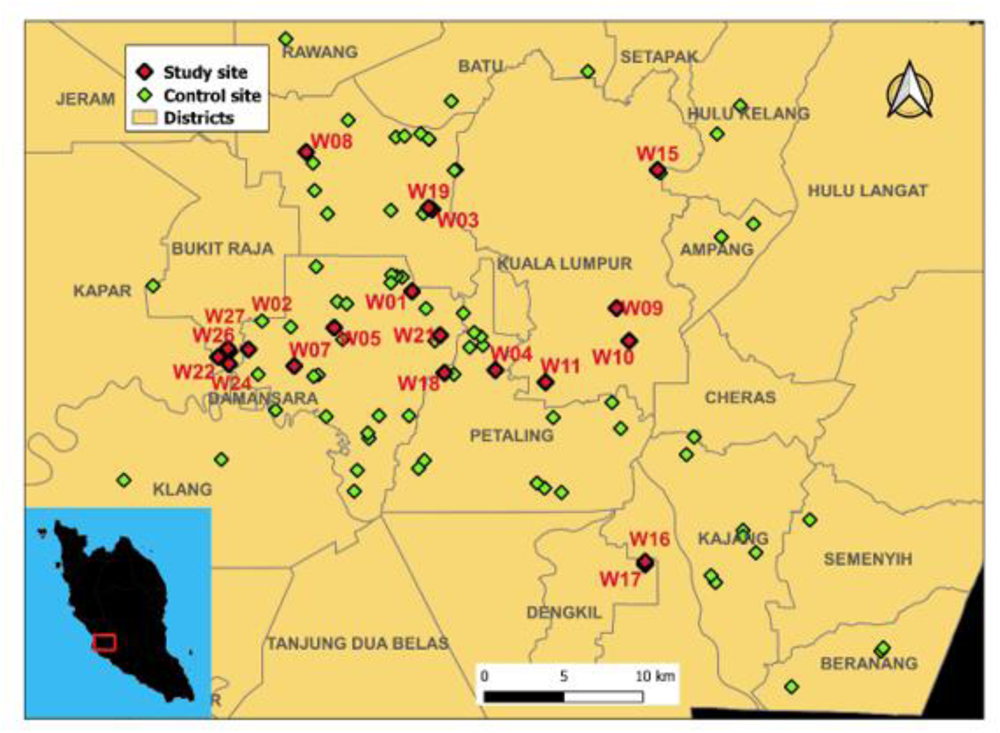
Map of Selangor region showing operational release sites used in evaluating *Wolbachia* impact and control sites. Districts within the region are labelled. Release sites are marked (see Table S1 for code).

Release sites (Table S1) consisted of areas that varied in size (48,692 to 276,261 m^2^) where *Ae. aegypti* numbers were relatively high (index value derived from the proportion of positive ovitraps placed for 7 days in an area > 40%). Sites with a high *Aedes aegypti* index and low *Aedes albopictus* index were preferred. For each defined area constituting a site, data on the incidence of dengue cases across the previous 5 years were collected, along with information on the relative abundance of *Ae. aegypti* and *Ae. albopictus* mosquitoes (based on prior ovitrap surveillance).

For each individual site nominated by authorities, a site profile was generated in terms of the number of residential housing blocks, number of stories, whether the area was treated with outdoor residual sprays and whether it was surrounded by any potential barriers to mosquito movement, particularly multi-lane highways or vegetated areas.

A fundamental requirement for successful implementation of a mosquito release program is community engagement involving comprehensive education of the local community. This ensures that the community appreciates the underlying objectives and strategies associated with the release initiative. The engagement efforts were undertaken by the Institute of Behavioural Research (IPTK) in collaboration with the Institute of Medical Research (IMR). Community members were regularly invited to IMR and provided with transparent updates of release progress. Community members were encouraged to be actively engaged in discussions to encourage shared responsibility and ownership. The local community were positive about the release of both male and female *Wolbachia* infected mosquitoes as a way of suppressing dengue and in many cases the mosquitoes encountered after releases were viewed as “good” mosquitoes with a reluctance to kill them even though continued local steps to control were encouraged by engagement staff. The significant reduction in dengue cases within release areas instilled a sense of protective custodianship which led to communities becoming strong advocates for the continuation of the *Wolbachia* program.

A novel communication tool involved the creation of a WhatsApp group to facilitate exchange of ideas, insights, and concerns within the community. Members sought clarifications, shared observations and engaged in dialogues that helped foster collective commitment to disease prevention. This contributed to vigilance in the community about pesticide fogging activities near to release areas communicated to local health departments which often led to the cessation of fogging operations. This represents a good example of community advocacy for the program. Community champions emerged which shared firsthand experience of release related activities and the insights of these community members extending to other release sites. In control sites community engagement consisted of larval source reduction activities.

Three days prior to the first release, a program of fogging, larval control, breeding site search and destroy activities, and health promotion activities were undertaken as outlined for previous releases [6]. These are normal procedures carried out in dengue hotspot areas as represented by the release and control sites assessed here. For the promotion activities, brochures on the benefits of releasing Aedes *Wolbachia* were generated and distributed to households.

### Operational releases

Eggs were initially used in the operational releases but there was a switch to adults later during the COVID -19 period (below). We aimed to release a total of 10 mosquitoes per house unit per week, with equal numbers of males and females being released. The number of eggs released was always double this number, with the expectation that 50% of the eggs would not hatch due for example to predation of eggs and adults emerging from egg release containers (see below).

For egg releases, 200 eggs were introduced into a release container as described in Nazni et al. [6]. Egg containers were placed in sites within buildings such as passageways and staircases to ensure minimum disturbance by the community, and in shaded spots to avoid direct exposure to sunlight, which can result in high larval mortality. Egg release containers were visited 3 times; on day 2 after placement to introduced food for larvae, on day 5 to open the stoppers or lids for adults to escape into the open, and on day 7 to collect the egg release containers and to replace these with another set of egg release containers.

The switch from eggs to adults related to the fact that the early release period coincided with COVID-19 pandemic, whereby the several phases of control on the human population were present. The Movement Control Order (MCO) was implemented, then the Conditional Movement Control Order (CMCO) and Recovery Movement Control Order (RMCO) until the end of May 2021. During the MCO, only key government and private services were allowed to operate, while the CMCO and RMCO aimed to revive the economy while implementing social distancing measures to effectively curb the progression of the pandemic. Standard operating procedures (SOPs) and guidelines consisting of health and safety measures was issued by the Malaysian National Security Council via mass media using radio, television, newspapers, government websites, social media, and text messages. During the MCO, the 3Cs (Crowded places, Confined spaces, Close conversation) and applying the 3Ws (Wash hands, Wear masks, Warn against risks, were strictly followed. Due to the introduction of MCO, egg releases were no longer possible and were changed to adult releases to reduce issues around communication and social distancing by MOH staff releasing the *Wolbachia*.

For adult releases, eggs were provided by IMR to the Botanic Laboratory Selangor (Ministry of Health) where eggs were hatched and reared to the adult stage, with releases involving 3-day-old adults (sexes mixed) held in small containers as described in Nazni et al. [6].

### Research sites

Monitoring continued at 5 of the release sites involving multistorey buildings described in Nazni et al [6]. This included *Wolbachia* monitoring yearly in Mentari Court, 6 monthly Flat A, B and C in Shah Alam, and 3-monthly in Section 7 Commercial Centre (see below).

### Control sites

The dengue confirmed cases for the high-rise hotspot localities in Malaysia were accumulated for five years periods since year 2014. The control sites were selected based on the localities with high occurrences of 5-years dengue outbreak in the Selangor district (Fig. 1). Seventy-six control sites were enlisted by the Vector Control Department. The control sites were selected to match operational and research sites in terms of involving multiple story residential buildings with a similar area size and population size (Table S1). However matched groupings had no impact on dengue reduction effects when included in the Bayesian models (below) so only analyses based on pooled control site data are presented.

### Wolbachia monitoring

Ovitraps were used to collect *Aedes* to assess *Wolbachia* frequencies. Each ovitrap consisted of a plastic container (96 mm height, 67 mm diameter) with 150 mL water and a wooden paddle (2 cm x 7 cm). In all *Wolbachia* release operational localities and research localities, 100 ovitraps were set up in the apartment buildings. The ovitraps were spread across every 2-3 floors. If there were carparks for the respective buildings, ovitrap were placed on alternate floors. Ovitraps were collected after a week and the paddle + water was transferred to a plastic container (12 x 3 x 12 cm). All emerging mosquitoes were identified to species and a maximum of 5 *Ae. aegypti* per trap were used for *Wolbachia* screening.

In all operational release sites, monitoring was done after 4 weeks from the first release. Adult releases and egg releases were stopped when frequency of *Wolbachia* in field population reached 80% and remained at 80% or more for 3 consecutive monitoring periods across 3 months. Monitoring was undertaken irregularly after that period while research sites were also irregularly monitored.

Adults from ovitraps were stored in absolute ethanol at − 80°C. DNA was extracted from individual mosquitoes using a glass bead and Chelex solution in an extraction tube. Mosquitoes were homogenized in 175 µL of 5% Chelex solution using TissueLyser II machine (QIAGEN) and with 5 µL of proteinase K (20 mg/mL) (Bioron Life Science). The extraction plate was spun down at 4000 rpm for 5 mins and the mosquito aliquot was transferred into 96 plates and spun down again for another 5 minutes. The extraction was incubated in a thermocycler at 65°C for 1 hour, followed by incubation for 10 min at 90°C. The plate was removed and spun down for another 5 mins. *Wolbachia* was detected by high-resolution melting polymerase chain reaction (qPCR-HRM) [27] with 1:10 diluted DNA using the following wAlbB1-specifc primers: wAlbB1-F (50-CCTTACCTCCTGCACAACAA) and wAlbB1-R (50 – GGATTGTCCAGTGGCCTTA), as well as universal mosquito primers: mRpS6_F (50-AGTTGAACGTATCGTTT CCCGCTAC) and mRpS6_R (5 0 - GAAGTGACGCAGCTTGTGGTCGTCC), which target the conserved region of the RpS6 gene, and *Ae. aegypti* primers aRpS6-F (5 0 -ATCAAGAAGCGCCGTGTCG) and aRpS6-R (5 0 -CAGGTGCAGGATCTTCATGTATTCG), which target the *Ae. aegypti*-specific polymorphisms within RpS6 and do not amplify *Ae. albopictus*.

Reactions were run as 384-well plates in a LightCycler 480 II (Roche). qPCR-HRM was performed in 10µL reactions containing 2 µL of DNA, 0.08 µL of 50 mM forward + reverse primer, 2.92 µL Milli-Q water and 5 µL Ronald’s Real-Time Buffer (3.28 µL Milli-Q water, 0.4 µL MgCl2 (50 mM), 1.0 µL ThermoPol Reaction Buffer with 20 mM Magnesium (10x), 0.25 µL HRM Master (Roche), 0.064 µL dNTPs (25 mM) and 0.01 µL Immolase (20 U/µL). qPCR was run following cycling conditions: 95°C for 10 min, followed by 50 cycles of 95°C for 10 s, 58°C for 15 s, 72°C for 15 s. High resolution melting was performed by heating the PCR product to 95°C, and then cooling to 40°C. Then the temperature was increased to 65°C. Samples were considered positive for *Wolbachia* when the Tm for the amplicon produced by the *Ae. aegypti* primers was at least 84°C and the Tm for the *Wolbachia*-primer amplicon was around 80°C.

### Dengue data

The dengue incidence for both intervention sites and control were obtained starting from 2013. Based on guidelines published by Disease Control Division, namely the Case Definition for Infectious Diseases in Malaysia (2017), a confirmed dengue case was defined as fulfilling both the clinical and laboratory criteria. Clinical criteria include acute onset of high-grade fever of 2-5 days associated with 2 or more clinical features such as headache, retro-orbital pain myalgia, arthralgia, rash and mild haemorrhagic manifestation. All patients presented with relevant symptoms and be diagnosed with suspected dengue will be notified by attending doctor and confirmatory laboratory test will be conducted. The laboratory criteria for dengue included detection of dengue non-structural protein 1 (NS1) or dengue antibody (IgM/IgG) or dengue virus genome by PCR or dengue virus isolation from serum or dengue virus antigen in tissue biopsy. Patients that tested positive for dengue with any of these tests are registered in national dengue registry, also known as eDengue. Dengue combo rapid test kits containing NS1 and IgM/IgG are used as point of care testing in most government health clinics for early diagnosis and treatment.

### Analysis of Wolbachia impact

A Bayesian time series model was used to estimate reduction in dengue cases as a result of the increased frequency of *Wolbachia* following releases. The model follows that in Nazni *et al.* [6], but the reduction in dengue cases is modelled as proportional to the frequency of *Wolbachia*. The model structure was as follows:

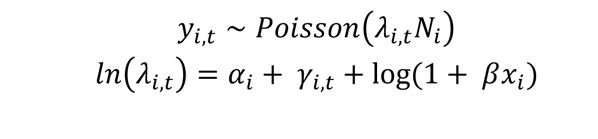

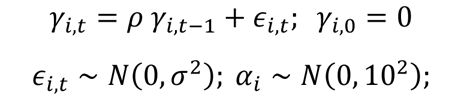

with global parameter prior distributions:

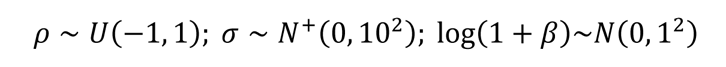

where the number of cases *y* at each site *i* and week *t* were assumed to follow a Poisson distribution, with the expected count given by the product of population at that site, and the per-capita incidence which varied varying through time and between sites. Each site had a separate time series of log-incidences *α*_*i*_ + *γ*_*i*,*t*_ with temporal correlation driven by an autoregressive model of order one, with parameters *ρ* and *σ*^2^shared by all sites. Each observation therefore had a separate temporally-correlated random effect on the log scale, to account for extra-Poisson dispersion and temporal correlation in case counts. Both release sites (including pre-and post-release periods) and non-release sites were included, enabling us to estimate expected levels of volatility in case count time series in this part of the model.

The intervention effect was represented by a parameter *β* and a covariate *x*_*i*,*t*_ giving the mean frequency of *Wolbachia* in the mosquito population in each site and time. The values of *x*_*i*,*t*_ were assumed to be constant over the release and post-release monitoring periods (set a the mean of observations), and set to 0 in pre-release times and in non-release sites. Model parameters were assigned vague priors with normal; positive-truncated normal; or uniform distributions. The log(1 + *x*) transformations applied to the intervention effect term ensures that the parameter *β* can be interpreted as the proportional change in dengue cases if *Wolbachia* frequency was at 100%, which is scaled linearly according to the achieved *Wolbachia* frequency. I.e. at a *Wolbachia* frequency of 100%, dengue cases are changed by percentage 100 ∗ *β*, whilst at a frequency of 25%, dengue cases are changed by percentage of 25 ∗ *β*. This choice of prior implies a median value of 0% change in dengue cases and a 50% interval ranging from a 49% reduction to a 96% increase in dengue cases. The model therefore assumes *a priori* that there is a 50% probability of *Wolbachia* presence reducing dengue cases, and a 50% probability of it increasing cases. The model was fitted to the dengue incidence data to estimate the impact of the releases, and to assess the evidence from the dengue case data that releases lead to a reduction in incidence - quantified as the posterior probability that *β* is negative.

Posterior samples of model parameters were simulated by Hamiltonian Monte Carlo in greta^29^ with 4 chains, each yielding 4000 posterior samples of model parameters after a warmup period of 1000 iterations during which period the leapfrog step size and diagonal mass matrix parameters were tuned. The number of leapfrog steps was sampled uniformly from between 30 and 40 throughout. Convergence was assessed by the Gelman-Rubin ^*R*^^ diagnostic, using the coda R package^30^ (^*R*^^ ≤ 1.01 for all parameters) and visual assessment of trace plots. Model fit was assessed by posterior predictive simulation: a random dataset of *y*_*i*,*t*_ values was generated according to each posterior sample of *ρ*_*i*,*t*_ and *r*, and the distributions of the simulated *y*_*i*,*t*_ values were compared with the observed *y*_*i*,*t*_. The analysis code is freely available online at https://github.com/goldingn/wolbachia_kl.

## Results

### Wolbachia invasion and persistence

Because releases were started in different areas at different times, changes in *Wolbachia* frequency are presented as weeks since the releases were initiated (Figure 2). The weeks when releases were undertaken are also presented on these graphs. *Wolbachia* frequencies increased rapidly at some sites and stayed high (W16, W17, W21, W24) which included two research sites. At others (W03, W05, W07, W10, W11, W19) they increased more slowly to reach a high frequency. At still others (W22, W27) there seemed to be a period of instability before frequencies settled at a high level. The remaining sites such as W02 and W15 showed instability which contributed to subsequent releases being undertaken (as marked on the graphs).

**Fig. 2.**
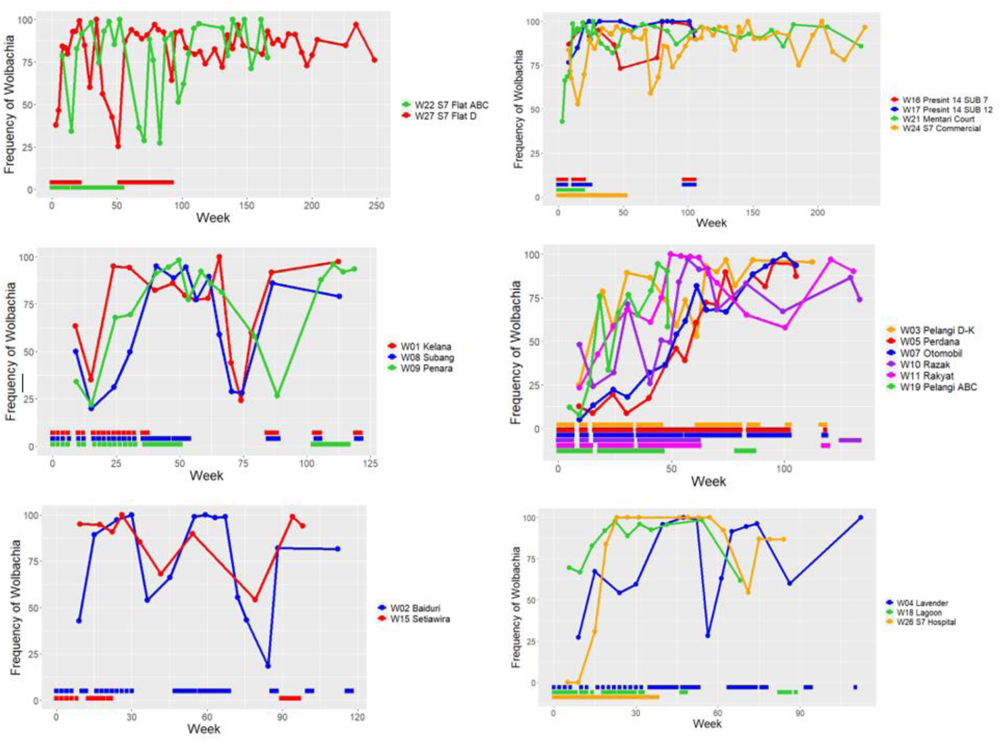
Changes in the frequency of *Wolbachia* across time in the release sites. The sites with similar patterns of *Wolbachia* changes are grouped together. Frequency estimates are based on a minimum of 10 adults being screened. Periods of mosquito release are also indicated on the graphs along the bottom colour coded for each release site.

### Dengue impact

Across all 20 release sites, the mean *Wolbachia* frequency in the post-release monitoring period averaged 82.3%, ranging from 56.2% to 96%. The posterior mean estimate of the reduction in dengue incidence at this average *Wolbachia* frequency was 62.4% (95% credible interval: 50-71% reduction), ranging from a 42.6% (95% CI: 34-49%) decrease in the lowest-frequency site to a 72.8% (95% credible interval: 59-83%) decrease in the highest frequency site. At a hypothetical 100% *Wolbachia* frequency, the model estimated a posterior mean reduction in dengue incidence of 75.8%, with a 95% credible interval from 61-87%. Table 1 summarises the posterior distributions over the model parameters. The posterior probability of *Wolbachia* releases resulting in a reduction in dengue incidence was greater than 0.999.

**Table 1.** Parameter estimates for variables in the model:

| Parameter | Posterior mean | 95% credible interval |
| --- | --- | --- |
| $\rho$ | 0.937 | 0.93 - 0.94 |
| $\sigma$ | 0.482 | 0.47 - 0.5 |
| $\beta$ | -0.758 | -0.87 - -0.61 |

**Figure 3.**
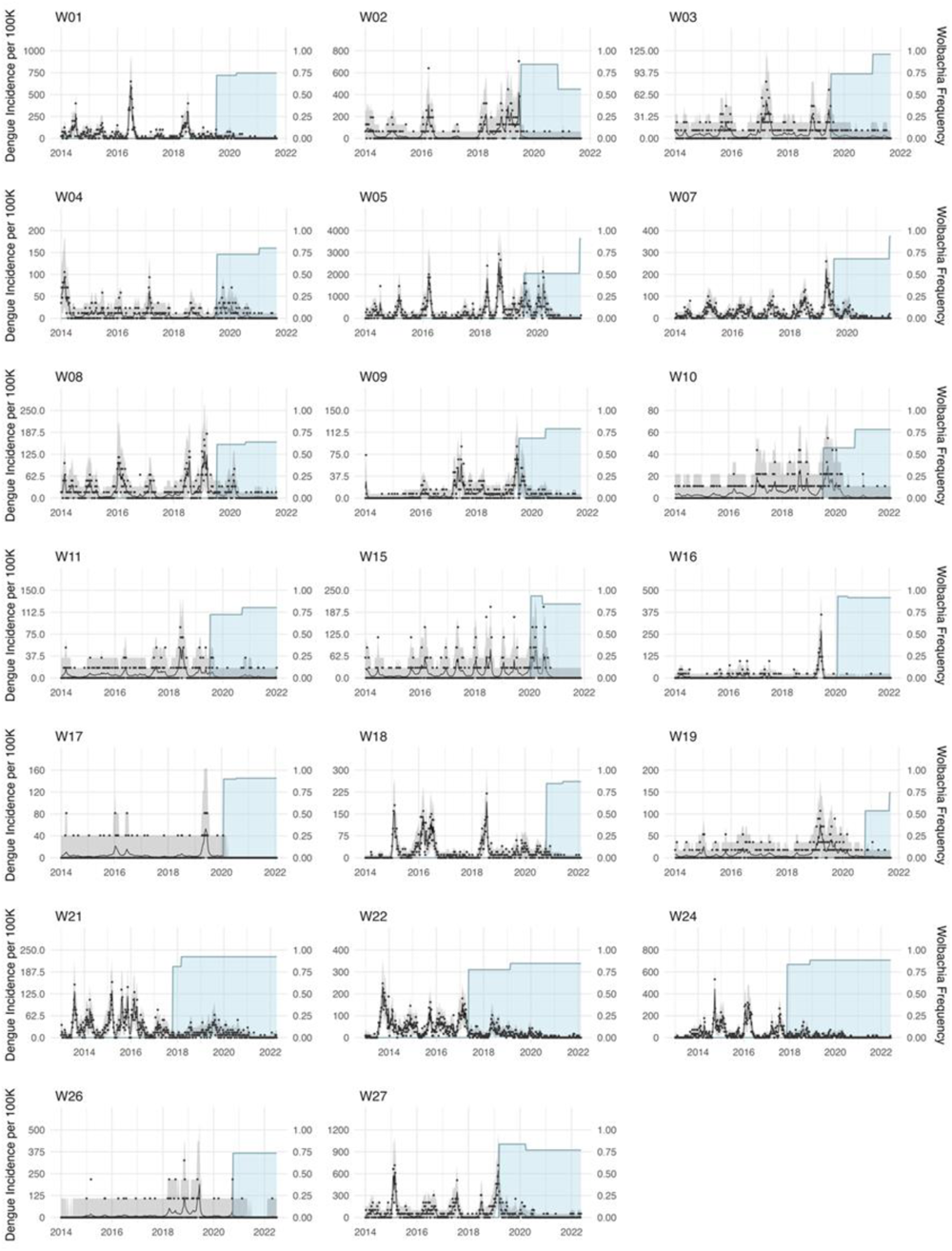
Incidence of dengue in the 20 release sites (data in black dots, posterior mean predicted incidence in black line, 95% predictive interval in grey) with the period of *Wolbachia* release and mean frequency of *Wolbachia* (blue region; first level is the release period, second is after releases finished).

## Discussion

With the additional release sites and extended period of dengue monitoring, we find evidence for stronger dengue reduction following release of *w*AlbB-carrying mosquitoes, particularly at high *Wolbachia* frequencies in release sites. At some research sites like Mentari Court and S7 Commercial, dengue cases have now been reduced for a number of years and the *Wolbachia* has remained stably high without further releases; in high rise blocks at Mentari Court, the *Wolbachia* also remains well spread throughout the apartment blocks where interventions took place [6, 16]. Dengue reduction has persisted despite no further community engagement in these sites. Clearly as *Wolbachia* frequencies stabilise in new areas there is the potential for further increasing the impact of the infection on dengue cases.

The level of dengue reduction is similar to that achieved in Yogyakarta with *w*Mel [8] although that project employed releases over larger areas in a randomised design rather than through an operationalized approach aimed at specific areas where dengue was common. *Wolbachia* frequencies in large, invaded areas are expected to be more stable than small areas since there is a lower likelihood of movement of *Wolbachia*-free mosquitoes into an area [18], although this will also depend on the structure of the urban environment including the presence of wide roads [19].

Dengue will be acquired outside release areas and contribute to the local incidence of dengue in an area, making it less likely to detect large reductions in small areas. Dengue reduction in areas where there have been lower *Wolbachia* frequencies recorded have been more moderate than in Yogyakarta [10] and confirm the direct association between dengue blocking *Wolbachia* and disease reduction.

The substantial impact of *Wolbachia* frequency on dengue reduction highlights the importance of continued *Wolbachia* monitoring and taking the necessary steps when there is a decrease in *Wolbachia* frequencies. Booster releases were successful in re-establishing or maintaining high frequencies (such as locations Kelana and Subang in Fig. 2) but this does entail ongoing monitoring of release sites. The reasons for the observed variation in *Wolbachia* stability remain unclear, with no obvious connection to any of the variables defining the release sites (Table 1).

However, it is consistent with data from release programs in other countries that highlight variable success in obtaining introductions which include slow increases in some populations [10, 20]. Identifying the factors involved remains an intriguing area for future research, with a number of factors that could be important such as environmental conditions, local mosquito movement patterns (including immigration from neighbouring areas such as construction sites with high mosquito density), and the nature of breeding sites (predominance of permanent versus temporary, periodically inundated breeding sites, which will affect *Wolbachia* fitness costs) [21–23]. Ongoing fogging in neighbourhoods adjacent to release sites is another possibility. Theory also emphasizes the importance of density dependent interactions [24]. A combination of local *Wolbachia* and entomological monitoring as well as molecular approaches such as tracking mtDNA variation [25] and new ways of tracking breeding sites [26] could provide useful tools in developing this understanding.

The outcome of this research programme followed by an expanded operational programme serves as the basis for future expansion of releases in additional dengue-prone areas. To date a total of 40 localities inclusive of research sites have been used for releases with *Ae. aegypti* carrying *w*AlbB *Wolbachia* in 8 states in Malaysia. The Malaysian Ministry of Health has established plans for future release of *Wolbachia* mosquitoes in dengue hotspots as a national rollout programme. The results support the continued use of *w*AlbB in the Kuala Lumpur area, where temperatures in breeding sites can be high, so the proven strong stability of *w*AlbB *Wolbachia* under hot conditions [27, 28] remains important.

## Data Availability

All data produced in the present study are available upon reasonable request to the authors

## Acknowledgments

Funding for the study was provided by Wellcome Trust Awards 226166, 108508, 202888 and Ministry of Health Malaysia NMRR-16-297-28898. The authors thank the Director General of Health, Ministry of Health, Malaysia, for permission to publish. We acknowledge Datuk Dr Norhayati binti Rusli (Deputy Director General of Health (Public Health), Ministry of Health, Datuk Dr Rose Nani Mudin, and Mohd Khairuddin Che Ibrahim for the continuous support throughout the project.

## Supplementary Information

**Supplementary Figure S1.**
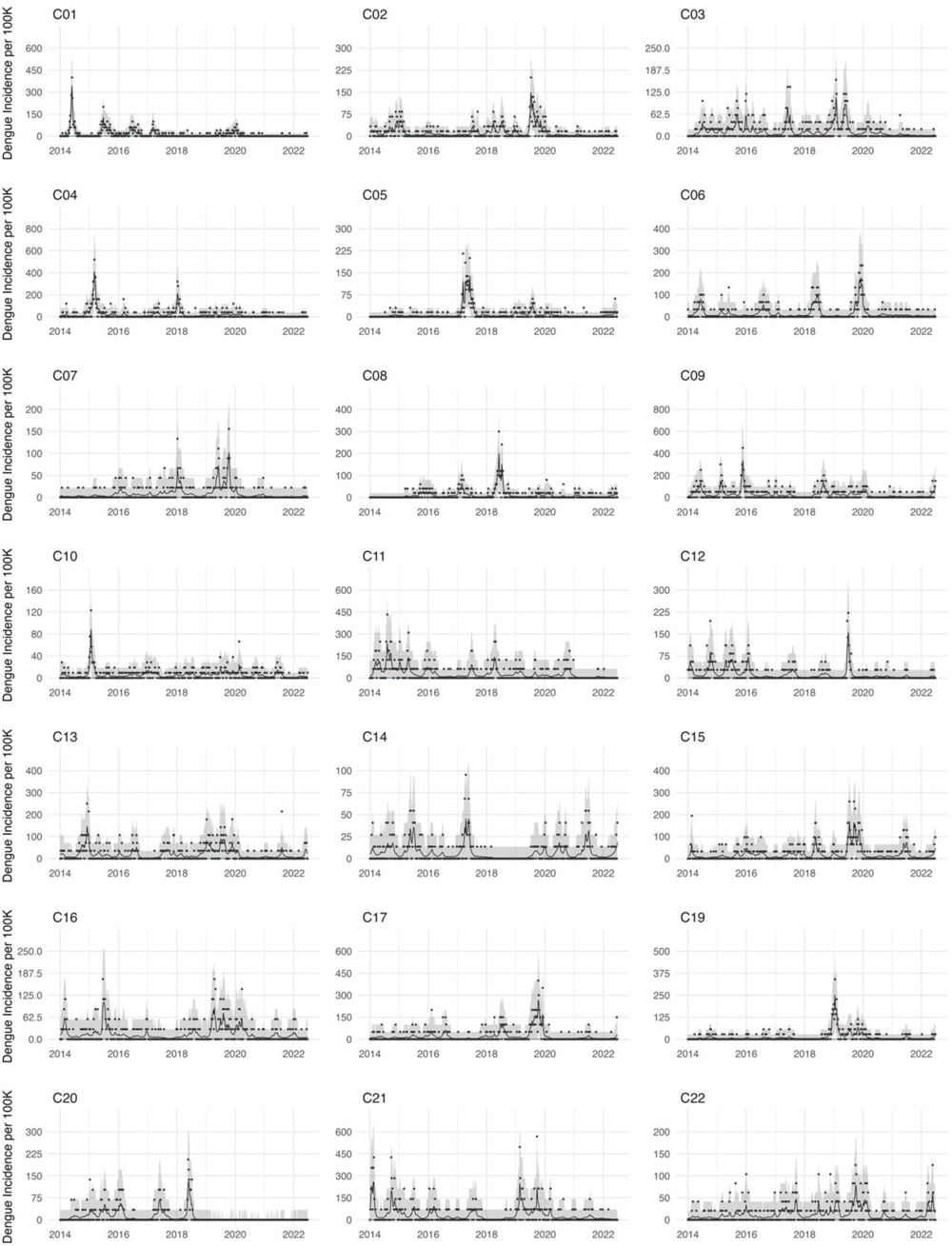
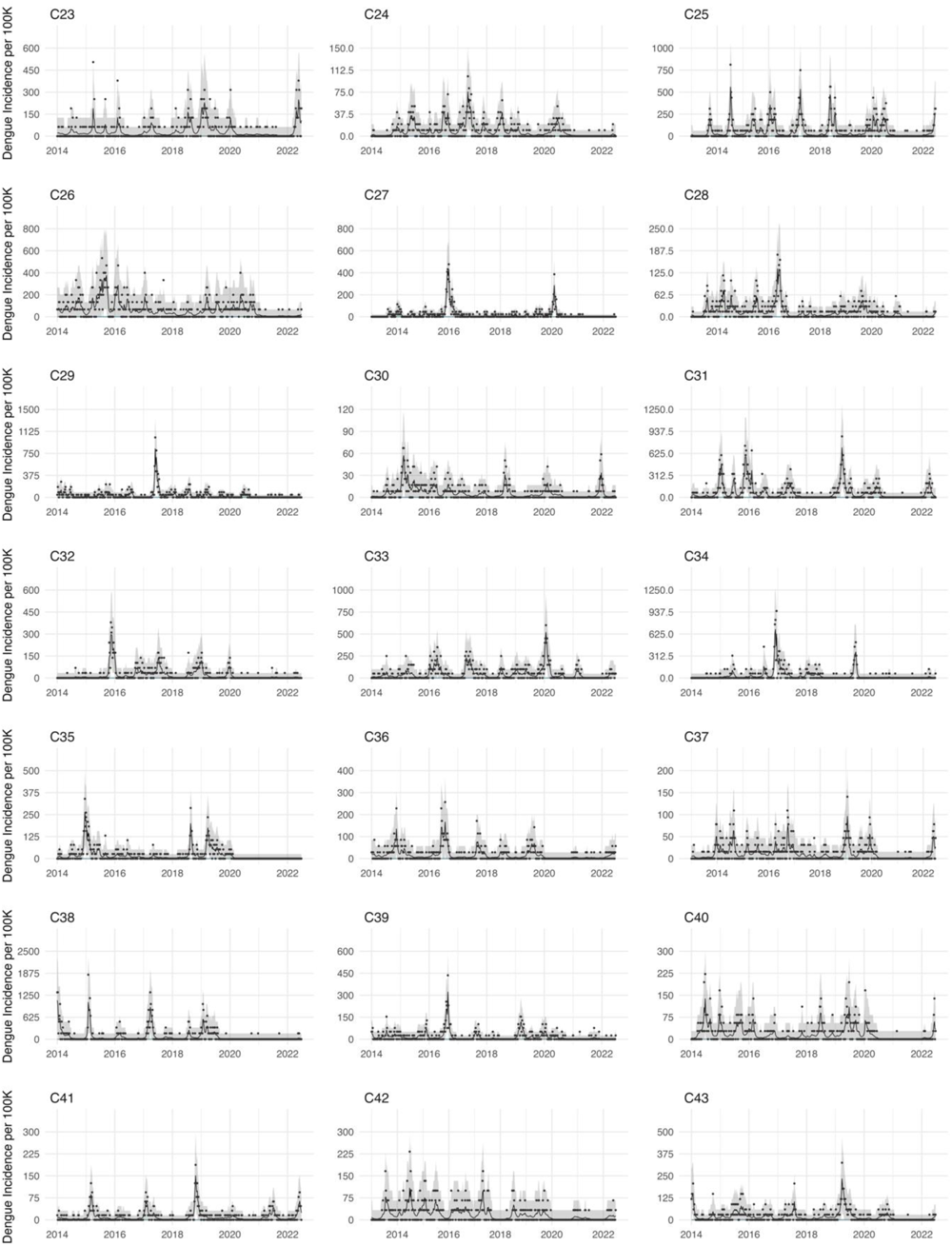
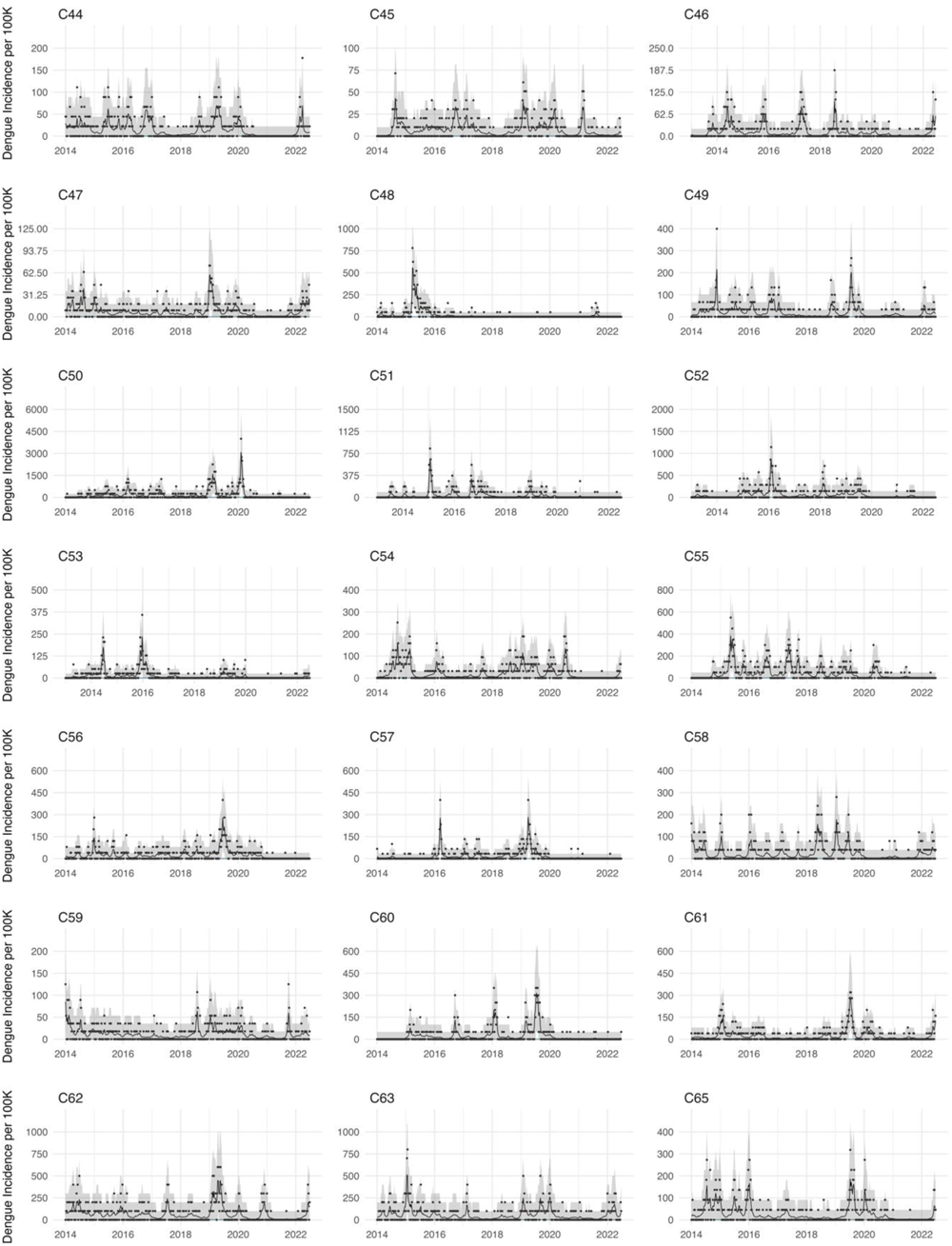
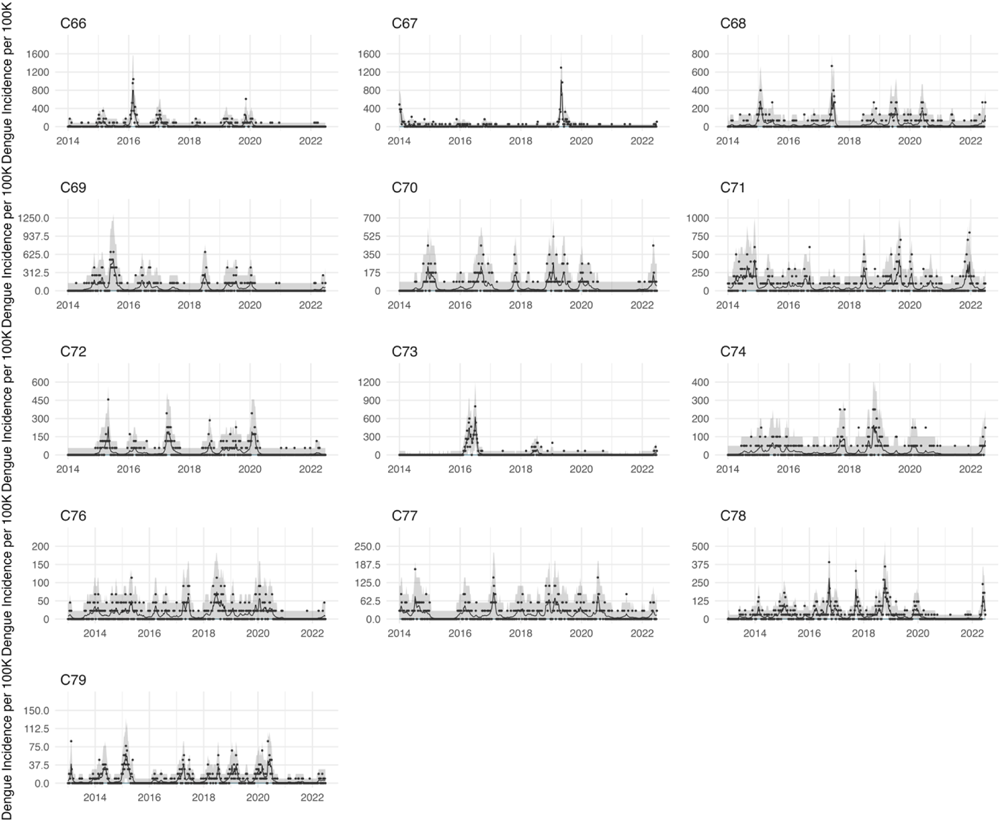
Incidence of dengue in 76 non-release sites (data in black dots, posterior mean predicted incidence in black line, 95% predictive interval in grey).

**Supplementary Table S1.** Details of release and control sites. Note that these are given as they were matched but in the analysis all control sites are considered together because there was no impact of matched group on dengue reduction.

| Site Code | Locality Name | Type of Site | Size (km<sup>2</sup>) | No of Level | Population | Starting Date |
| --- | --- | --- | --- | --- | --- | --- |
| W01 | SS7 Kelana Putera/Puteri Condo | Study site | 0.08694 | 18 | 4000 | 8-Jul-19 |
| C35 | Kd Pju 1a Pprt A: Blok A,B,C | Control site | 0.00891 | 18 | 3825 |  |
| C74 | Tm Pjs 2d Flat Desa Mentari (Blok 11) | Control site | 0.0177298 | 18 | 2000 |  |
| C31 | Seksyen U2 (Pangsapuri Ilham) | Control site | 0.02285 | 18 | 1500 |  |
| W02 | Seksyen 7 Apartment Baiduri | Study site | 0.021 | 11 | 1560 | 10-Jul-19 |
| C38 | Seksyen U2 (Pangsapuri Jaya) | Control site | 0.0193693 | 5 | 600 |  |
| C50 | Tmn Puchong Permai (Flat Blok F) | Control site | 0.001369 | 5 | 400 |  |
| C09 | Appt Seri Cempaka Seksyen 16 Bandar Baru Bangi | Control site | 0.041073 | 5 | 2000 |  |
| C57 | Seksyen U19 (Rkat Kem Paya Jaras) | Control site | 0.3125071 | 5 | 3000 |  |
| C56 | Tmn Puchong Permai (Flat 38 Blok A) | Control site | 0.0829443 | 5 | 2500 |  |
| W03 | BI PJU 6 Pelangi Damansara | Study site | 0.0489 | 20 | 8640 | 9-Jul-19 |
| C03 | Sri Maya Condo | Control site | 0.5 | 20 | 5,000 |  |
| C43 | Sri Pjs 10 Ridzuan Kondo | Control site | 0.027 | 28 | 3388 |  |
| C73 | Subang Jaya (Dk Senza Condominium Jln Taylors) | Control site | 0.0161982 | 23 | 1500 |  |
| W04 | Puchong Seksyen 3 Apartment Vista Lavender | Study site | 0.06296 | 12 | 8545 | 9-Jul-19 |
| C44 | Kd Pju 5 Seksyen 4 Gugusan Semarak | Control site | 0.0562 | 5 | 4500 |  |
| C15 | Blok N-X Pangsapuri Baiduri Bandar Tasik Kesuma | Control site | 0.01 | 5 | 3080 |  |
| C22 | B.B.T Flat Jalan Batu Nilam 34 | Control site | 0.0837 | 5 | 4818 |  |
| C49 | BI Pju 10 Apt Harmoni 2 (D,E,F,G,H,J,K,T,S,R ) | Control site | 0.0495 | 5 | 3000 |  |
| C32 | Seksyen U13 (Pangsapuri Seroja ) | Control site | 0.0520632 | 6 | 2900 |  |
| W05 | Seksyen 13 Apartment Perdana | Study site | 0.025446 | 5 and 10 | 750 | 8-Jul-19 |
| C70 | Seksyen U5 (Pangsapuri Mutiara Subang : Blok 1 - 7 ) | Control site | 0.0561095 | 5 | 1150 |  |
| C21 | Appt Sri Anggerik Seksyen 7 Bandar Baru Bangi | Control site | 0.049165 | 5 | 1408 |  |
| C65 | BI Pju 9 Sd 13 Apt Sri Meranti: Blok A - G | Control site | 0.1181917 | 5 | 2200 |  |
| C17 | Blok A-G Pangsapuri Baiduri Bandar Tasik Kesuma | Control site | 0.01 | 4 | 2000 |  |
| C13 | Appt Sri Tanjung Seksyen 7 Bandar Baru Bangi | Control site | 0.1 | 5 | 2800 |  |
| W07 | Seksyen 15 Dataran Otomobil | Study site | 0.06191 | 5 | 10000 | 7-Jul-19 |
| C14 | Appt Taman Setia Balakong | Control site | 0.18 | 5 | 7348 |  |
| C08 | Pangsapuri Nuri Bandar Bukit Mahkota | Control site | 0.91 | 4 | 5000 |  |
| C26 | Tmn Equine (Flat Putra Permai & Rumah Kedai ) | Control site | 0.094 | 5 | 1500 |  |
| C06 | Appt Casmara | Control site | 0.1 | 5 | 3,000 |  |
| C40 | BI Pju 9 Sd 13 Apt Sri Meranti 2: Blok H, J, K, L, M, N, P, Q, R | Control site | 0.04396 | 5 | 3600 |  |
| W08 | Seksyen U5 Pangsapuri Subang Suria | Study site | 0.078789 | 7 | 6000 | 10-Jul-19 |
| C02 | Appt Palma Bch | Control site | 0.2 | 5 | 6,000 |  |
| C07 | Appt Sri Putri Up | Control site | 0.2 | 5 | 4,500 |  |
| C72 | Usj 15 (Subang Perdana Goodyear Court 10a) Blok Aa-Aq | Control site | 0.0723245 | 5 | 1750 |  |
| C16 | Appt Taman Prima Cempaka | Control site | 0.036342 | 5 | 3510 |  |
| W09 | Cheras Pangsapuri Sri Penara | Study site | 0.111 | 17 | 13575 | 7-Jul-19 |
| C47 | Tm Pjs 2c Apt Medan Cahaya (Blok A - H) | Control site | 0.0654722 | 16 | 11000 |  |
| C10 | Flat Sri Nilam Bandar Baru Ampang | Control site | 0.04 | 16 | 10560 |  |
| C01 | Appt Residensi Bistaria | Control site | 0.4 | 15 | 5,000 |  |
| W10 | Cheras PPR Desa Tun Razak | Study site | 0.041 | 21 | 9120 | 8-Jul-19 |
| C62 | Kd Pju 3 Palm Spring Kondo 2: Blok D, E, F | Control site | 0.0424987 | 21 to 24 | 4360 |  |
| C46 | Tmn Pinggiran Putra (Vista Pinggiran) | Control site | 0.02593 | 20 | 4800 |  |
| C41 | Usj 16 (Sri Tanjung Appt) | Control site | 0.03825 | 10 | 6400 |  |
| W11 | Bukit Jalil Apartment Sri Rakyat | Study site | 0.043 | 16 | 5760 | 7-Jul-19 |
| C05 | Appt Laguna Biru | Control site | 0.3 | 18 | 6,500 |  |
| C33 | Seksyen 26 (Flat Pprt: Blok A, B, C ) | Control site | 0.009923 | 17 | 2000 |  |
| C23 | Appartment Golden Villa | Control site | 0.03 | 6-15 | 1584 |  |
| W15 | RKAT Desa Setia Wira, Setiawangsa | Study site | 0.07 | 16 | 3450 | 16-Jan-20 |
| C77 | TM Sek 51A Flat Impian Baiduri | Control site | 0.0206896 | 17 | 3500 |  |
| C29 | Tmn Serdang Perdana (South City Condo) | Control site | 0.0052448 | 16 | 2250 |  |
| C61 | Kd Pju 1a Pprt B: Blok D,E,F | Control site | 0.0094356 | 17 | 2500 |  |
| C60 | Putra Height 9 (Jln Putra Indah - Sri Mutiara Appt) | Control site | 0.0287637 | 10 | 2000 |  |
| W16 | Presint 14, Parcel sub 12 (kuarters kerajaan) | Study site | 0.06 | 19 | 4150 | 23-Jan-20 |
| C19 | Flat Desa Lembah Permai | Control site | 0.5 | 11 | 3520 |  |
| C67 | Seksyen 27 (Apt Ixora) | Control site | 0.037556 | 5-11 | 1850 |  |
| C12 | Appt Taman Impian Indah | Control site | 0.04 | 5 | 3600 |  |
| W17 | Presint 14, Parcel sub 7 (kuarters kerajaan) | Study site | 0.037 | 17 | 2450 | 23-Jan-20 |
| C63 | Tm Pjs 2b Flat Desa Mentari (Blok 9, 10) | Control site | 0.0283855 | 18 | 1000 |  |
| C68 | Kd Pju 1a Pprt C: Blok G, H | Control site | 0.0083678 | 17 | 1500 |  |
| C54 | Seksyen 9 (Kondo Sri Mahligai & Town House Sri Mahligai ) | Control site | 0.05727 | 3 to 13 | 3170 |  |
| W18 | PJS 9 Pangsapuri Lagoon Perdana | Study site | 0.0618705 | 18 | 10000 | 12-Oct-20 |
| C59 | Sri Pjs 5 Flat Desa Ria | Control site | 0.0541775 | 5-15 | 5600 |  |
| C55 | Tmn Serdang Perdana Psrn Perdana (One South) | Control site | 0.0562384 | 16 | 2000 |  |
| C39 | Sri Ss 6 Kompleks Kediaman Kastam | Control site | 0.06762 | 14 | 3900 |  |
| W19 | Pelangi Damansara Blok A-C | Study site | 0.02155 | 19 | 5600 | 15-Oct-20 |
| C48 | Kd Pju 1a Puncak Sri Kelana Kondo | Control site | 0.02563 | 15 | 1920 |  |
| C04 | Appt Desa 1 | Control site | 0.35 | 10 to 12 | 2500 |  |
| C45 | Bdr Kinrara 6f (Enggang Apt) | Control site | 0.03967 | 10 | 9800 |  |
| C30 | Usj 1 (Angsana Appt) | Control site | 0.029581 | 13 | 11900 |  |
| C24 | Tmn Serdang Perdana Sp 5 (Sky Villa, Kesidang & Rumah Kedai) | Control site | 0.06075 | 20 | 9800 |  |
| C71 | BI Pju 10 Apt Lestari | Control site | 0.0575238 | 10 | 1000 |  |
| W21 | Mentari Court | Study site | 0.090267 | 17 | 13876 | 16-Oct-17 |
| C28 | BI Pju 8 Apt Flora Damansara 2 (Blok C - F) | Control site | 0.00343 | 24 | 6800 |  |
| C42 | BI Pju 8 Apt Flora Damansara 1 (Blok A, B) | Control site | 0.007858 | 24 | 3010 |  |
| C25 | Seksyen 13 (Pangsapuri Brunsville Riverview) | Control site | 0.0106505 | 12 | 1600 |  |
| W22 | Section 7 Flats (Flat A, B & C) | Study site | 0.276261 | 5 | 11720 | 13-May-17 |
| C79c | Seksyen U3 (Pangsapuri Daisy A: Blok 1-2, B: Blok 3-8, C : Blok 9-13) | Control site | 0.095803 | 5 | 10400 |  |
| C76b | Seksyen 8 (Flat A&B : Blok 1-22) | Control site | 0.1627001 | 5 | 4400 |  |
| C78a | Seksyen 16 (Flat A: Blok 1-4 & Blok 9-16, Flat B: Blok 5-8, Flat C: Blok 17-26 & Pangsapuri Anggerik Indah) | Control site | 0.054822 | 4 and 10 | 3328 |  |
| C53 | Seksyen 20 (Flat B: Blok 13 - 19) | Control site | 0.04099 | 5 | 3900 |  |
| W24 | Section 7 Commercial Centre | Study site | 0.119025 | 4 and 5 | 5632 | 20-Nov-17 |
| C27 | Seksyen 27 (Flat A: Blok 1 - 22 ) | Control site | 0.0841 | 5 | 4400 |  |
| C52 | Seksyen U5 (Pangsapuri Orkid: Blok 7 - 12) | Control site | 0.0093324 | 5 | 700 |  |
| C37 | Tmn Lestari Perdana (Pangsapuri Sri Indah) | Control site | 0.089613 | 5 | 6400 |  |
| W26 | Section 7 Hospital Quarters | Study site | 0.019921 | 11 | 920 | 29-Sep-20 |
| C66 | Seksyen 28 Flat A: Blok 1, 3, 5, 7, 9, 11, 13, 15, 17, 19) | Control site | 0.0317079 | 4 | 1150 |  |
| C34 | Seksyen U7 (Tudm Subang D: Blok F1 - F8 & E1 - E2) | Control site | 0.0209267 | 5 | 1575 |  |
| C69 | BI Pju 10 Indah Kondo | Control site | 0.0491161 | 5 | 750 |  |
| W27 | Section 7 Flat D | Study site |  | 5 | 1960 | 11-Mar-19 |
| C36 | BI Pju 10 Apt Permai | Control site | 0.089731 | 5 | 3500 |  |
| C51 | Seksyen 24 (Flat J: Blok 79 - 84) | Control site | 0.02628 | 5 | 1080 |  |
| C58 | Seksyen 20 (Kuarters Polis: Blok 5 - 12 ) | Control site | 0.0486504 | 5 | 2500 |  |
| C11 | Appt Taman Tenaga | Control site | 0.26 | 5 | 1614 |  |
| C20 | Appt Jalan 6c/9-6c/11 Seksyen 16<br>Bandar Baru Bangi | Control site | 0.17 | 5 | 2916 |  |

